# Neighbourhood built environments, socioeconomic position, and hospital admissions for cardiovascular disease: a prospective study using UK Biobank

**DOI:** 10.1101/2023.05.22.23289600

**Authors:** Kate Mason, Neil Pearce, Steven Cummins

## Abstract

**Background:** Neighbourhood environments may influence cardiovascular disease (CVD) risk, e.g. by influencing diet and physical activity (PA) behaviours. We explored whether associations between characteristics of neighbourhood environments and CVD are modified by area deprivation and household income. If effects of neighbourhood risk exposures vary by socioeconomic position, efforts to improve population health by improving neighbourhood built environments could widen health inequalities.

**Methods:** In the UK Biobank cohort we used linked records of hospital admissions to assess the relative hazard of being admitted to hospital with a primary diagnosis of CVD according to three characteristics of the neighbourhood built environment: availability of formal PA facilities, proximity of a takeaway/fast-food store, and neighbourhood greenspace. We then examined potential effect modification of the main associations by household income and area deprivation. We used Cox proportional hazards models, adjusted for likely confounding, and calculated relative excess risks due to interaction (RERI) to assess effect modification on the additive scale. We also examined the combined modifying role of income and deprivation.

**Results:** There were 13,809 incident CVD admissions in the sample (mean follow-up=6.8 years). Overall associations between neighbourhood exposures and CVD-related hospital admissions were weak to null. However, there was evidence of effect modification by both area deprivation and household income. Greater availability of PA facilities near home was associated with lower risk of CVD-related admission in more deprived areas, but only among people in higher-income households. Area deprivation and household income both modified the association with fast-food proximity. More greenspace was not associated with lower risk of CVD-related admission for any group. Some results differed between women and men. Findings were largely robust to alternative model specifications.

**Conclusions:** Improving deprived neighbourhoods by increasing the number of PA facilities, while also ensuring access to these is free or affordable, may improve population health. Examining effect modification by multiple socioeconomic indicators in parallel can yield deeper insight into how different aspects of the people’s socioeconomic conditions influence their relationship with the built environment and its effects on their health. Improved understanding may help to avoid generating or perpetuating health inequalities when neighbourhood-based built environment interventions are designed.

## BACKGROUND

Inadequate physical activity (PA), poor diet and chronic stress are risk factors for cardiovascular disease^1–3^. Features of neighbourhood environments have the potential to either support or hamper healthy diet and PA behaviours, and to mitigate or exacerbate the stresses of daily life. Unequal access to healthy neighbourhood resources may therefore result in differential risk of cardiovascular disease.

Over the past 25 years, cross-sectional studies have produced inconsistent evidence linking neighbourhood built environment characteristics to cardiovascular health^4, 5^ (and similarly to adiposity and obesity^6^, mental health^7^, and health behaviours such as PA and diet^8, 9^). Recently, findings from longitudinal studies have also contributed to the evidence base (e.g.^10, 11^), and by providing greater certainty about the temporal sequence of exposures and outcomes of interest, these study designs may help to better elucidate the true causal relationships between neighbourhood environments and these outcomes. Linkage of hospital records and mortality registers to population-based cohorts with geographical data on neighbourhood environments provides opportunities to examine whether environmental characteristics of neighbourhoods are associated with objectively recorded, prospective outcomes, consistent with hypothesised relationships.

Numerous features of the neighbourhood built environment are widely hypothesised to influence cardiovascular health, via either diet or PA. These include, but are not limited to, the retail food environment (including proximity or density of healthy and unhealthy food stores), accessibility of recreation facilities for physical activity (such as public swimming pools, gyms, sports fields), and green spaces such as public parks and private gardens (which may facilitate recreational PA, or functional PA such as gardening or active travel). Greenspace may also offer additional health benefits via other pathways relating to the regulation of stress hormones, improved immune function through exposure to diverse microorganisms, and reduced exposure to air pollution^12^, all of which may influence risk of cardiovascular disease and other chronic diseases. The evidence base for the effect of neighbourhood exposures on cardiovascular health remains inconclusive, and the relative importance of different neighbourhood exposures is unclear. Any causal effects of neighbourhood exposures on health are likely to be small, and bound up in a larger set of environmental, social and structural drivers of health behaviours and outcomes that collectively contributing to the complex physical and social contexts within which we make lifestyle choices and navigate the modern world.

Understanding these relationships requires us to recognise that they may not be uniform across the population, but rather, certain individuals or groups in specific locations may be more sensitive to the influences of their local environment than others. Average, population-wide estimates might mask important heterogenous effects – those that are concentrated in particular population subgroups or particular places. One likely source of such effect heterogeneity is socioeconomic position. Results of some studies suggest differential neighbourhood health effects by individual socioeconomic status^13, 14^ or neighbourhood deprivation^15, 16^. These may arise if the availability of particular neighbourhood resources is different in more deprived areas compared with less deprived areas^17^, or if there are differential preferences for using particular neighbourhood resources according to individual socioeconomic status, regardless of availability e.g. if low-income households frequent fast-food/takeaway stores more often, or differential access e.g. unaffordable membership fees for gyms and leisure centres. In the case of greenspace effects on health, differences may arise according to area-level deprivation (rather than individual/household socioeconomic position) if more deprived areas have poorer quality greenspace. On the other hand, if greenspace access is free to the individual, then access may offset inequitable access to formal PA facilities, and therefore have a greater effect for low-income households. Whatever the direction of any such effect heterogeneity, this remains a poorly understood aspect of the relationships between neighbourhood environments and health. If differential benefits or harms of neighbourhood characteristics do exist along individual and/or area socioeconomic axes, there is a risk that efforts to improve population health by improving neighbourhood built environments may widen health inequalities if socially differential impacts are ignored^18^.

In this study we use baseline UK Biobank data on neighbourhood exposures to PA facilities, fast-food stores and greenspace, linked to records of hospital admissions up to January 2016, to examine (1) the relative hazard of being admitted to hospital with a primary diagnosis of cardiovascular disease, according to exposure to each of the neighbourhood characteristics, and (2) whether there is evidence of effect modification of those associations by household income and/or area deprivation.

## METHODS

### Study design and data collection

We used data from UK Biobank (project 17380), the scientific rationale, study design and survey methods for which have been described elsewhere^19^. All individuals aged 40–69 years listed on National Health Service (NHS) patient registers and living within a 25-mile radius of any of the study’s 22 assessment centres across the UK were invited to participate in the study. Roughly half a million individuals were recruited to visit a UK Biobank assessment centre between 2006 and 2010. The 40-69 year age range was selected by UK Biobank as an important period for the development of many chronic diseases.

### Linked hospital admissions data

UK Biobank features ongoing prospective linkage of the cohort to administrative health records. At the time of analysis, linked Hospital Episode Statistics were available up to January 2016. These contain information on hospital admissions coded using the International Statistical Classification of Diseases and Related Health Problems, 10th Revision (ICD-10). We used these data to identify incident admissions to hospital for cardiovascular disease.

### Outcomes

Our outcome for this study was any hospital admission for which the primary diagnosis is recorded as cardiovascular disease, namely ICD-10 codes I10-I25, I46,I48,I50,and I60-79.

### Neighbourhood environment data

Also linked to UK Biobank is a high-resolution spatial database of objectively measured characteristics of the physical environment surrounding each participant’s exact residential address, known as the UK Biobank Urban Morphometric Platform (UKBUMP). Environmental data in UKBUMP were derived, using automated processes, from multiple pre-existing sources roughly contemporaneous with the individual baseline assessment^20^. Subsequently, additional environmental measures have been linked to the cohort, including the measure of greenspace^21^ that we have used here in addition to two measures from the original UKBUMP.

### Neighbourhood exposures

Three measures of the neighbourhood built environment were examined. To account for skewed distributions and to facilitate a categorical approach to the analysis of effect modification^22^, each exposure was split into four categories. The exposures we examined were:

(1) Availability of PA facilities: number of formal PA facilities within a one-kilometre street-network distance of each participant’s home address, categorised as 0, 1, 2-3, or 4 or more.
(2) Fast-food proximity: street-network distance in metres from participants’ home address to the nearest ‘hot/cold fast-food outlet/takeaway’, categorised as <500 m, 500-999 m, 1000-1999 m, 2000 m+.
(3) Greenspace: percentage of 300 m Euclidean buffer around home address classified as ‘greenspace’ or ‘domestic garden’ in the Generalised Land Use Database. Combining ‘greenspace’ and ‘gardens’ is consistent with previous research using the GLUD to examine relationships with health^23^. A 300 m buffer was chosen to capture greenspace in the immediate vicinity of a person’s home. There is some evidence that 300 m is a distance from home beyond which the use of green spaces quickly declines^24, 25^, and it has been proposed in the UK as a benchmark for greenspace provision^26^. Greenspace was grouped into quartiles.

Exposures (1) and (2) were derived in the UKBUMP from OS AddressBase Premium 2012^20^, while (3) was derived by Wheeler et al from the Generalised Land Use Database 2005^21^. We restricted the analyses to people residing in England, because the greenspace data for exposure (3) were not available for UK Biobank participants in Wales and Scotland.

### Potential effect modifiers

We examined whether the association between each neighbourhood exposure and CVD admissions was modified by binary indicators for annual, pre-tax household income (<£31,000 or ≥£31,000) and area deprivation (most deprived 40% of UK census output areas vs least deprived 60%, based on the Townsend index). When testing for effect modification, household income and area deprivation were combined with each primary exposure into a categorical variable capturing all combinations of levels of the exposure and potential modifier, with a single reference category (see below for details). We also examined the combined modifying role of income and deprivation. Area-based and individual indicators of socioeconomic disadvantage have been shown to contribute to health outcomes independently of one another, providing a rationale for examining them both in parallel and in combination^27^.

### Potential confounders

We identified potential confounders of the primary associations as age (years), sex (binary), ethnicity (White/non-White), educational qualifications (College or University degree/A levels/AS levels or equivalent/O levels or below/other), employment status (paid work, retired, unable to work, unemployed, or other), urban/non-urban status, UK Biobank assessment centre, and neighbourhood residential density (count of residential dwellings within a 1-km street-network buffer of home address, log transformed). Annual household income (<£18,000, £18,000–30,999, £31,000–51,999, at least £52,000) and area deprivation (Townsend score) were also included as possible confounders in any models where they were not being tested as a potential effect modifier. We also adjusted models for smoking status (current/previous/never), alcohol intake frequency (less than/at least 3 times per week) as these are important risk factors for CVD and may be correlated with neighbourhood, and number of years living at current (baseline) address to at least partially condition on pre-baseline exposure to neighbourhood environment, which could act as a confounder.

### Statistical analysis

Of the 502,617 participants in UK Biobank for whom some data were available, 502,544 remained after withdrawals were excluded, and 355,691 of these individuals lived in England and had complete data on covariates and data for at least one measure of the neighbourhood environment. Of these, we excluded 19,535 individuals who had reported prior cardiovascular events from analyses involving CVD outcomes. This left a possible N= 336,156 for the analysis (Figure 1). The final analytic sample sizes varied according to the neighbourhood variable under examination. The maximum follow-up time after baseline assessment was 9.8 years, but varied according to the date of an individual’s recruitment to the study.

**Figure 1.**
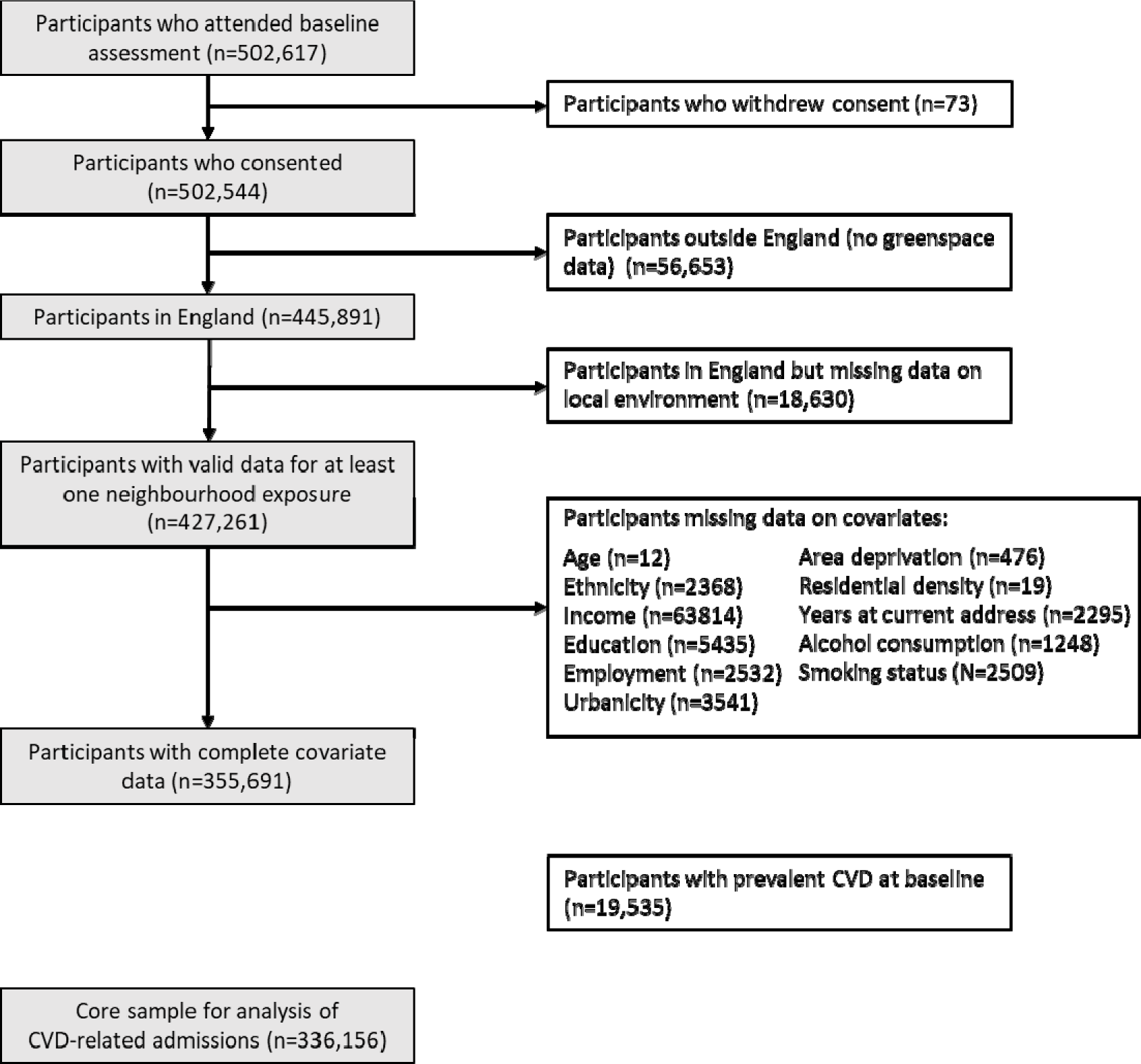
UK Biobank sample for analyses.

Baseline characteristics were summarised by the mean (and standard deviation) or median (and interquartile range) for continuous variables and number (and percent) for categorical variables. We then examined associations between neighbourhood exposure and incident hospital admission due to CVD following baseline assessment using multivariable Cox proportional hazard models, with adjustment for potential confounders and censoring for death. Results are expressed as hazard ratios (HRs) and 95% confidence intervals (95% CI). The proportional hazards assumption was tested by visual inspection of adjusted log-log plots (Supplementary Figure 2). The reference categories for each neighbourhood exposure are the hypothetically most health-damaging (lowest availability of PA facilities, shortest distance to nearest fast-food store, least greenspace).

We examined whether the primary associations were modified by area deprivation and household income. In line with STROBE recommendations^28^ and using the method described by Li and Chambless^29^ and VanderWeele^22^, the relative excess risk due to interaction (RERI) was calculated to assess effect modification on the additive scale. When dealing with binary and time-to-event outcomes, the decision to examine effect modification on either the multiplicative or the additive scale has implications for interpretation. The additive scale provides important information about the potential public health consequence of intervening on the exposure, for different strata of the effect modifier. This is not information we can glean directly from an examination of effect modification on the multiplicative scale, because measures of effect modification on the multiplicative scale ignore potentially different baseline risks within strata of the effect modifier^22^. The RERI is calculated by estimating the HR for each combination of the exposure and potential modifier values relative to a single reference category, in this case the least hypothetically health-promoting level of the respective neighbourhood variable (no PA facilities; <500 m from nearest fast-food store; or quartile with least greenspace), and either low income (<£31,000) or more deprived area (home address located in a census output area in the most deprived 40% of all UK areas). In other words, the reference category in each analysis is the group expected to have the highest baseline risk of the outcome. From this model, the RERI is calculated as:

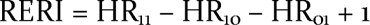

For the model assessing effect modification of PA facility availability by household income, for example, HR_11_ represents the HR (relative to the reference category) for people in high-income households (at least £31,000 per year) and who have the highest level of neighbourhood availability of PA facilities (4 or more within 1000 m of home); HR_10_ represents the HR for people in low-income households with 4 or more PA facilities near home; and HR_01_ is the HR for people in high-income households with no PA facilities near home.

For the models of the other neighbourhood exposures, and models of effect modification by area deprivation, subscript *1* represents those most exposed to the potentially health-promoting neighbourhood exposure and less deprived areas, respectively. As such, a RERI value greater than zero – which implies a positive departure from additivity – suggests that in this case any estimated protective effect of the neighbourhood variable among people in low-income households or in more deprived areas is greater than the estimated protective effect among people from high-income households or less deprived areas. In contrast, a RERI<0 suggests any protective effect of the neighbourhood variable is greater in the high-income/less deprived group. All analyses were conducted using Stata v14.2 (StataCorp LP, College Station, TX, USA).

### Sensitivity analyses

The spatial data used in the creation of the UKBUMP to ascertain the neighbourhood food and physical activity exposures were recorded in 2012, just after the baseline data collection period^20^, and while it is assumed that neighbourhood exposure will be sufficiently constant over this period as to not unduly influence the results, we check this assumption by conducting a sensitivity analysis in which follow-up is restricted to the period from 2012 onwards for all participants, rather than from the baseline assessment date (which could be as early as 2006).

The primary analyses were not adjusted for baseline hypertension or BMI, or medications for hypertension or cholesterol, because of ambiguity regarding temporal precedence. Although these are important risk factors for CVD, rather than being confounders they may be on the causal pathways from neighbourhood environment to CVD if neighbourhood exposure predates them, which it is likely to in this largely residentially stable population (median time living at baseline address was 16 years). We instead conduct sensitivity analyses to examine whether adjusting for these variables influences the primary findings. We also report the main associations stratified by sex.

## RESULTS

### Descriptive

Table 1 summarises the characteristics of the study participants at the baseline assessment. The analytic sample has a mean age of 56 years at baseline and was predominantly of White ethnicity and urban dwelling. Reflecting the age of the sample, just over half were educated to no higher than O levels, and six in every ten were employed at baseline. Participants were evenly distributed across income categories, with roughly half living in households with an annual gross income below £31,000, while 29% lived in the more deprived 40% of areas in the UK.

**Table 1.** Descriptive characteristics of sample (N=336,156)

| *Variable* | *Participants*<br>*admitted for CVD*<br>*(n=13,809)* | *Participants not*<br>*admitted for CVD*<br>*(n=322,347)* | *Total sample*<br>*(n=336,156)* |
| --- | --- | --- | --- |
| % female | 33.97% | 54.80% | 53.94% |
| Age (mean, SD) | 60.29 (6.69) | 55.75 (8.07) | 55.94 (8.07) |
| Ethnicity (% non-White) | 4.11% | 4.97% | 4.94% |
| % Urban | 86.09% | 85.45% | 85.48% |
| Education (%) |  |  |  |
| College or University degree | 27.97% | 35.25% | 34.95% |
| A levels/AS levels or equivalent | 9.84% | 11.88% | 11.80% |
| O levels or below/other | 62.19% | 52.87% | 53.26% |
| Employment status |  |  |  |
| Paid work | 45.98% | 62.75% | 62.06% |
| Retired | 45.99% | 29.62% | 30.29% |
| Unable to work | 4.62% | 2.35% | 2.44% |
| Unemployed | 1.55% | 1.53% | 1.53% |
| Other | 1.87% | 3.76% | 3.68% |
| Residential density (residential sites per 1000m<br>buffer) (median, IQR) | 1932<br>(1118–3057) | 1914<br>(1101–3133) | 1915<br>(1102–3129) |
| Years at current address (median, IQR) | 19 (9–28) | 15 (7–25) | 15 (7–25) |
| Area deprivation (mean Townsend score) | -1.32 (3.03) | -1.46 (2.94) | -1.46 (2.95) |
| Area deprivation (% in two most deprived<br>quintiles of the UK) | 30.29% | 28.80% | 28.86% |
| Household income |  |  |  |
| <£18,000 | 30.37% | 21.10% | 21.48% |
| £18 000–30 999 | 28.89% | 25.32% | 25.46% |
| £31 000–51 999 | 23.11% | 26.71% | 26.56% |
| £52 000 or more | 17.63% | 26.88% | 26.50% |
| Smoking status |  |  |  |
| Current | 14.28% | 9.95% | 10.13% |
| Previous | 41.05% | 34.14% | 34.42% |
| Never | 44.66% | 55.91% | 55.45% |
| Frequency of alcohol consumption (% ≥3 times<br>per week) | 45.92% | 45.42% | 45.44% |

The mean follow-up time for participants was 6.8 years. Over the follow-up period, 13,809 individuals (4.11%) were admitted to hospital with CVD (Table 2). Proportionally, there were more hospital admissions for CVD among people from low-income households, whereas admissions were similar across levels of area deprivation.

**Table 2.** Hospital admissions by household income and area deprivation.

|  | *Total N* | *Number of CVD admissions (%)* |
| --- | --- | --- |
| Total | 336,156 | 13,809 (4.11) |
| Household income (annual pre-tax) |  |  |
| <£31,000 | 157,818 | 8185 (8.19) |
| £31,000 or more | 178,338 | 5624 (3.16) |
| Area deprivation |  |  |
| More deprived | 97,005 | 4183 (4.31) |
| Less deprived | 239,151 | 9626 (4.03) |

\* Self-reported average total household income before tax
\*\* 'More deprived' refers to people living in areas in the top two most deprived quintiles of the UK, based on the Townsend index.

### Associations between neighbourhood characteristics and CVD admissions

Figure 2 summarises the hazard ratios for hospital admissions due to CVD associated with each of the three neighbourhood environment measures, across the sample as a whole. While 95% CIs for all associations included the null value of 1.0, there was some indication of a weak trend toward decreasing hazard of CVD-related hospital admission with increasing distance to the nearest takeaway/fast-food store. With each category decrease in proximity, the HR for hospital admission moved further away from 1.0, and those living further from a fast-food store had a 4% reduced hazard compared with those living closest (HR=0.96; 95%CI: 0.91-1.02). For neighbourhood availability of PA facilities and greenspace, there was little to no evidence of an association with risk of CVD-related admission when averaging across the whole study population.

**Figure 2.**
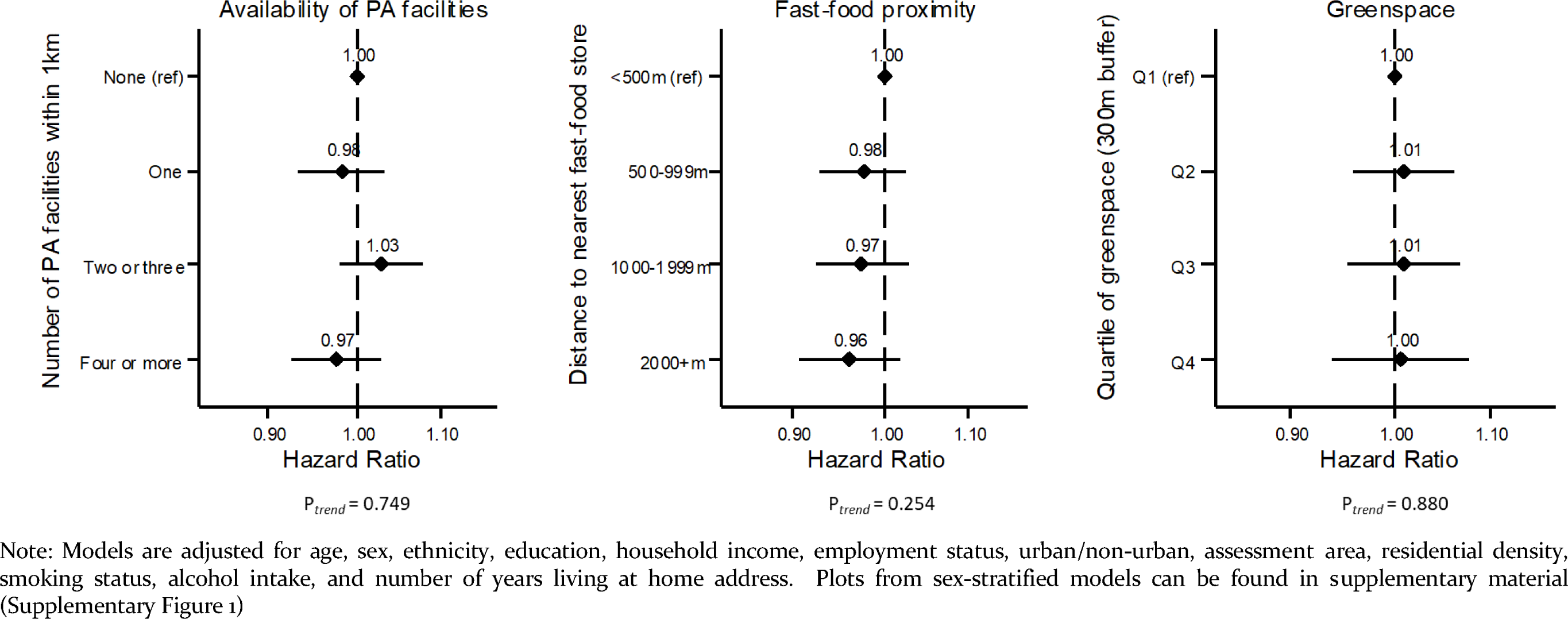
Adjusted hazard ratios for hospital admission due to CVD, by availability of formal PA facilities, proximity to nearest fast-food/takeaway store, and neighbourhood greenspace.

### Modification of the associations between neighbourhood characteristics and CVD-related hospital admissions, by income and area deprivation

Table 3 shows results of the analyses of effect modification by household income and area deprivation, for the associations between each neighbourhood exposure and CVD-related hospital admissions. For availability of PA facilities within a kilometre of home, there was some evidence of effect modification by area deprivation on the additive scale, with a RERI of 0.088 indicating a departure from additivity and a stronger association among people from more deprived areas. Among those living in areas in the two quintiles of greatest deprivation, the stratum-specific hazard ratio for those with at least four PA facilities near home was 0.90 (95%CI: 0.82–0.99), while we observed no association among people in less deprived areas (HR=1.01; 95%CI: 0.95–1.07), suggesting a greater protective effect in more deprived areas. The RERI for effect modification by household income, whilst not as large (RERI=-0.077), indicated a negative departure from additivity, suggesting effect modification in the other direction and a stronger association between PA facilities and CVD among those from higher income households. In stratified models the hazard ratio for those in higher income households with at least four PA facilities near home compared with none was 0.93 (95%CI: 0.85–1.00), while no such association was observed among people in lower income households (HR=1.00; 95%CI: 0.94–1.07).

**Table 3.** Modification of the association between built environment variables and hospital admissions due to CVD, by household income and area deprivation

| *CVD-related admissions* | *Annual household income\** |  | *Area deprivation\*\** |  |
| --- | --- | --- | --- | --- |
|  | < £31,000 | At least £31,000 | More deprived | Less deprived |
|  | *HR (95% CI)* | *HR (95% CI)* | *HR (95% CI)* | *HR (95% CI)* |
| **Number of PA facilities** |  |  |  |  |
| None (ref) | 1.00 | 0.99 (0.92, 1.05)<br>P=0.666 | 1.00 | 0.88 (0.81, 0.95)<br>P=0.001 |
| One | 1.00 (0.94, 1.07)<br>P=0.956 | 0.94 (0.87, 1.01)<br>P=0.086 | 0.93 (0.84, 1.03)<br>P=0.152 | 0.88 (0.81, 0.95)<br>P=0.002 |
| 2-3 | 1.04 (0.98, 1.10)<br>P=0.214 | 0.99 (0.92, 1.07)<br>P=0.834 | 0.98 (0.90, 1.07)<br>P=0.696 | 0.92 (0.85, 1.00)<br>P=0.040 |
| 4 or more | 1.00 (0.94, 1.07)<br>P=0.886 | 0.92 (0.85, 0.98)<br>P=0.015 | 0.92 (0.84, 1.00)<br>P=0.051 | 0.88 (0.81, 0.96)<br>P=0.004 |
| Stratum-specific HRs<br>(4+ facilities vs 0) | 1.00 (0.94, 1.07)<br>P=0.957 | 0.93 (0.85, 1.00)<br>P=0.062 | 0.90 (0.82, 0.99)<br>P=0.038 | 1.01 (0.95, 1.07)<br>P=0.821 |
| Relative excess risk due to<br>interaction (RERI) (95% CI) | -0.077 (-0.167, 0.013) P=0.092 |  | 0.088 (-0.004, 0.181) P=0.061 |  |
| **Fast-food proximity** |  |  |  |  |
| Closer than 500m (ref) | 1.00 | 0.92 (0.84, 1.00)<br>P=0.041 | 1.00 | 0.98 (0.91, 1.06)<br>P=0.615 |
| 500-999m | 0.97 (0.91, 1.03)<br>P=0.353 | 0.90 (0.84, 0.98)<br>P=0.009 | 1.01 (0.94, 1.09)<br>P=0.771 | 0.92 (0.86, 0.99)<br>P=0.026 |
| 1000-1999m | 0.97 (0.91, 1.03)<br>P=0.308 | 0.91 (0.84, 0.98)<br>P=0.012 | 0.97 (0.90, 1.06)<br>P=0.548 | 0.93 (0.87, 1.00)<br>P=0.055 |
| At least 2000m | 0.93 (0.87, 1.00)<br>P=0.048 | 0.92 (0.85, 1.00)<br>P=0.046 | 1.03 (0.93, 1.14)<br>P=0.595 | 0.90 (0.84, 0.98)<br>P=0.011 |
| Stratum-specific HRs<br>(≥2000m vs <500m) | 0.93 (0.86, 1.00)<br>P=0.047 | 1.01 (0.92, 1.11)<br>P=0.854 | 1.03 (0.93, 1.16)<br>P=0.547 | 0.92 (0.85, 0.99)<br>P=0.019 |
| Relative excess risk due to<br>interaction (RERI) (95% CI) | 0.076 (-0.020, 0.171) P=0.122 |  | -0.104 (-0.225, 0.017) P=0.093 |  |
| **Greenspace** |  |  |  |  |
| Q1 (least greenspace) (ref) | 1.00 | 0.91 (0.84, 0.98)<br>P=0.015 | 1.00 | 0.98 (0.91, 1.05)<br>P=0.547 |
| Q2 | 1.00 (0.94, 1.06)<br>P=0.897 | 0.94 (0.87, 1.01)<br>P=0.094 | 1.02 (0.95, 1.10)<br>P=0.560 | 0.94 (0.88, 1.00)<br>P=0.070 |
| Q3 | 0.99 (0.93, 1.06)<br>P=0.790 | 0.95 (0.88, 1.02)<br>P=0.154 | 1.05 (0.95, 1.15)<br>P=0.340 | 0.93 (0.87, 1.00)<br>P=0.036 |
| Q4 (most greenspace) | 0.98 (0.90, 1.05)<br>P=0.529 | 0.95 (0.88, 1.04)<br>P=0.275 | 0.98 (0.87, 1.11)<br>P=0.761 | 0.94 (0.87, 1.01)<br>P=0.104 |
| Stratum-specific HRs<br>(Q4 vs Q1) | 0.99 (0.91, 1.08)<br>P=0.844 | 1.02 (0.91, 1.13)<br>P=0.747 | 0.96 (0.83, 1.12)<br>P=0.605 | 0.96 (0.88, 1.04)<br>P=0.295 |
| Relative excess risk due to<br>interaction (RERI) (95% CI) | 0.070 (-0.023, 0.164) P=0.142 |  | -0.019 (-0.154, 0.115) P=0.778 |  |

\* Self-reported average total household income before tax. \*\* 'More deprived' refers to people living in areas in the top two most deprived quintiles of the UK, based on the Townsend index. Q= quartile
Note: RERI>0 indicates positive effect modification and a departure from additivity. Here, because the reference category is people with the hypothetically least-healthy level of neighbourhood exposure and with low household income or living in more deprived areas, a positive departure from additivity suggests any estimated *protective* effect of the neighbourhood variable is *weaker* among people in higher income households or in less deprived areas than it is among people from low income households or more deprived areas, and therefore that the latter stand to gain more from an intervention. In contrast, a RERI<0 suggests any protective effect of the neighbourhood variable is stronger in the high-income/less deprived group.

For fast-food proximity, we observed effect modification by both household income (RERI=0.076) and area deprivation (RERI=-0.104), but in the opposite direction from what was observed for PA facilities (Table 3). Reduced access to fast-food/takeaway stores might have the biggest impact for low-income households rather than higher income households, and mostly in less deprived areas. In stratified models, for people in low-income households, living at least 2 km from a fast-food store was associated with a 7% reduction in the hazard of CVD-related hospital admission compared with living within 500 m of a fast-food store (HR-0.93; 95% CI: 0.86-1.00), while no such association was observed among higher income households (HR=1.01; 95% CI: 0.92-1.11). But for people living in more deprived areas, there was no observed protection afforded by living further from a fast-food store, whereas for those in more affluent areas, living at least 2km from a store was associated with an 8% reduced hazard of hospitalisation for CVD (HR=0.92; 95% CI: 0.85-0.99).

Household income and area deprivation did not appear to modify associations between greenspace and CVD-related hospital admissions. There was some weak evidence of effect modification by income (RERI=0.070), but the stratum-specific HRs were null in both income groups (Table 3).

Combining area deprivation and household income, the hazard ratio for at least four PA facilities (compared with none) is smallest among people from high-income households living in more deprived areas (Table 4). Among this group, the hazard of being hospitalised for CVD was 22% lower for people who had at least four PA facilities within a kilometre of their home, compared with having no nearby PA facilities (HR=0.78, 95% CI: 0.65-0.94).

For fast-food proximity, a beneficial association of living further from a fast-food/takeaway store was only observed among low-income households in affluent areas, where the hazard of CVD-related admission was 12% lower among people living ≥2km from a fast-food store than among people living <500 m from one (HR=0.88, 95%CI:0.80-0.97; Table 4).

**Table 4.** Association between neighbourhood characteristics and CVD-related hospital admissions, stratified by household income and area deprivation in combination.

|  | *Combined household income and area deprivation* |  |  |  |
| --- | --- | --- | --- | --- |
|  | **Less than £31,000**<br>**& more deprived** | **At least £31,000**<br>**& more deprived** | **Less than £31,000**<br>**& less deprived** | **At least £31,000**<br>**& less deprived** |
|  | *HR (95% CI)* | *HR (95% CI)* | *HR (95% CI)* | *HR (95% CI)* |
| **Number of PA facilities** |  |  |  |  |
| None (ref) | 1.00 | 1.00 | 1.00 | 1.00 |
| One | 0.96 (0.85, 1.08)<br>P=0.461 | 0.84 (0.69, 1.03)<br>P=0.103 | 1.02 (0.95, 1.10)<br>P=0.574 | 0.96 (0.88, 1.05)<br>P=0.372 |
| 2-3 | 1.00 (0.90, 1.11)<br>P=0.977 | 0.90 (0.75, 1.08)<br>P=0.267 | 1.06 (0.98, 1.14)<br>P=0.144 | 1.01 (0.93, 1.10)<br>P=0.745 |
| 4 or more | 0.95 (0.85, 1.07)<br>P=0.398 | 0.78 (0.65, 0.94)<br>P=0.010 | 1.03 (0.95, 1.12)<br>P=0.481 | 0.96 (0.88, 1.05)<br>P=0.390 |
| *P<sub>trend</sub>* | 0.543 | 0.022 | 0.300 | 0.620 |
| **Fast-food proximity** |  |  |  |  |
| Closer than 500m (ref) | 1.00 | 1.00 | 1.00 | 1.00 |
| 500-999m | 1.03 (0.94, 1.13)<br>P=0.547 | 0.99 (0.85, 1.14)<br>P=0.872 | 0.93 (0.85, 1.01)<br>P=0.085 | 0.97 (0.88, 1.08)<br>P=0.613 |
| 1000-1999m | 0.96 (0.87, 1.07)<br>P=0.481 | 1.04 (0.88, 1.23)<br>P=0.670 | 0.95 (0.87, 1.04)<br>P=0.298 | 0.96 (0.87, 1.07)<br>P=0.454 |
| At least 2000m | 1.00 (0.88, 1.14)<br>P=0.969 | 1.12 (0.90, 1.39)<br>P=0.301 | 0.88 (0.80, 0.97)<br>P=0.013 | 0.98 (0.88, 1.09)<br>P=0.724 |
| *P<sub>trend</sub>* | 0.700 | 0.332 | 0.042 | 0.780 |
| **Greenspace** |  |  |  |  |
| Q1 (least greenspace) (ref) | 1.00 | 1.00 | 1.00 | 1.00 |
| Q2 | 1.01 (0.92, 1.11)<br>P=0.798 | 1.10 (0.95, 1.28)<br>P=0.210 | 0.99 (0.90, 1.08)<br>P=0.803 | 0.96 (0.86, 1.07)<br>P=0.467 |
| Q3 | 1.05 (0.93, 1.17)<br>P=0.428 | 1.06 (0.86, 1.29)<br>P=0.600 | 0.97 (0.88, 1.07)<br>P=0.550 | 0.98 (0.88, 1.10)<br>P=0.728 |
| Q4 (most greenspace) | 0.93 (0.78, 1.12)<br>P=0.449 | 1.08 (0.81, 1.44)<br>P=0.591 | 0.98 (0.88, 1.10)<br>P=0.786 | 0.99 (0.87, 1.12)<br>P=0.832 |
| *P<sub>trend</sub>* | 0.954 | 0.436 | 0.713 | 0.936 |

\* Self-reported average total household income before tax
\*\* 'More deprived' refers to people living in areas in the top two most deprived quintiles of the UK, based on the Townsend index.
Q = quartile

For neighbourhood greenspace, confidence intervals around all HRs were too wide to conclude that there was a CVD-related benefit of more greenspace in any of the income/deprivation combinations (Table 4).

### Sex differences

Examining sex differences in the primary results, we found that for the whole population, an association between the formal PA environment and CVD admissions was observed among women but not men (Supplementary Table 1a). No effect modification was observed for women. The lack of association among men, however, obscured socioeconomic differences: men in deprived areas and men in higher income households had reduced hazard of CVD-related hospital admission when they had greater neighbourhood availability of PA facilities.

For fast-food proximity and CVD-related admissions, the primary results help for men, but no associations were observed for women, for any socioeconomic group (Supplementary Table 1b). The lack of evidence for any association between neighbourhood greenspace and CVD-related admissions, across all socioeconomic strata, was consistent for men and women (Supplementary Table 1c).

### Sensitivity analyses

In general, restricting follow-up to the period from 2012 onwards for all participants, rather than from the baseline assessment date, reduced precision around point estimates, but made minimal difference to the overall direction and magnitude of most coefficients and RERI estimates (Supplementary Tables 3 & 4). The main exception to this was that the departure from additivity due to area deprivation for the association between fast-food proximity and CVD-related admissions was far less when follow-up was restricted (RERI=-0.058 cf. RERI=– 0.104). The RERI for effect modification by area deprivation of the association between PA facilities and CVD was also somewhat attenuated (RERI=0.070, cf. RERI=0.088), but in contrast effect modification of the same relationship by household income was amplified (RERI=-0.113 cf. RERI=-0.077), and the overall finding for the combination of income and deprivation was, if anything, stronger when follow-up was restricted to 2012 onwards. Sensitivity analyses adjusting for baseline BMI, hypertension and medications for cholesterol and hypertension, yielded very similar point estimates to the primary results (Supplementary Tables 5 & 6).

## DISCUSSION

Across this very large sample of adults in mid-life in the UK, we examined the relationship between three characteristics of the neighbourhood built environment and hospital admissions due to CVD, over almost 10 years of follow up, and whether these associations were modified by area deprivation and household income, with the aim of identifying potential targets for intervention to improve health without widening health inequalities.

For the overall sample, we observed at most a weak trend of a reduced hazard of hospital admission due to CVD with increasing distance to the nearest takeaway/fast-food store, and some protection for people with the greatest availability of PA facilities within one kilometre of home; however, the 95% confidence intervals did not exclude the null value of no hazard reduction. No association was apparent between neighbourhood greenspace coverage and CVD.

More noteworthy, however, are the findings for effect modification by household income and area deprivation, where we observed some interesting patterns that may help to illuminate important elements of the links between the neighbourhood built environment and health. The largely null associations overall appeared to be masking potentially important variation in the strength and magnitude of some of those associations according to socioeconomic conditions, especially when viewing the results for area deprivation and household income together.

For the availability of formal PA facilities and CVD outcomes, we observed some evidence that this relationship varied by area deprivation, suggesting that intervening to improve access to formal PA facilities in deprived areas is likely to have a greater public health impact than it would in less deprived areas. At the same time, our examination of effect modification by household income suggests the association was stronger among higher-income households (after controlling for area deprivation). Although the 95% CIs for both RERIs include zero, the direction of the RERI for income is consistent with the fact that most formal PA facilities impose some financial cost on users; i.e. if they are not free to access, we would expect health benefits of greater neighbourhood availability of these facilities to accrue disproportionally to higher-income households. The contrasting directions of the additive measures of effect modification by (higher) income and (less) deprivation imply that greater availability of formal PA facilities is particularly beneficial in deprived areas, but only for the those who can afford access. Consistent with these findings analysing the two socioeconomic measures separately, when we then considered household income and area deprivation together, the estimated benefits of greater availability of neighbourhood PA facilities was indeed largely restricted to high-income households in deprived areas, among whom we observed a one-fifth reduction in the hazard of being admitted to hospital with CVD for people living near at least four PA facilities, compared with those people who had no local PA facilities. If the CVD-related benefits of greater availability of PA facilities accrue primarily to high-income households in deprived areas, the policy implications are obvious: locating more PA facilities in deprived areas may reduce CVD risk, but the greatest gains stand to be made if facilities in those areas are accessible to all, regardless of income. Otherwise, health benefits in deprived areas may be concentrated among the well-off living there, thus widening health inequalities. A recent quasi-experimental study in a deprived local authority in the north west of England^30^ showed improved outcomes following the introduction of universal free access to council leisure facilities, indicating that this is indeed a potentially effective approach to adopt.

With respect to fast-food proximity and risk of CVD-related hospital admission, again the largely null main associations appear to conceal stronger associations in some subgroups. The findings for effect modification by income and area deprivation on the additive scale suggest that reducing access to fast-food stores may have the greatest impact for low-income households, especially men in those households, but mostly in relatively affluent areas. The RERI for area deprivation as a modifier of the association with CVD is less than zero, and stratified analysis shows that greater distances from home to the nearest fast-food store are more strongly associated with reduced risk of CVD-related hospital admissions among people living in less deprived areas. This may imply a greater range of alternatives to fast food available to residents of more affluent areas. If so, this supports other evidence for the importance of considering the whole retail food environment – healthy and unhealthy stores – when making planning decisions. And as was seen for PA facilities, there was evidence of a contrasting role of income: but in this case the impact of fast-food proximity seems to be stronger among people in low-income households. The latter finding is consistent with as similar recent study using UK Biobank but concentrating only on participants in London^14^ and makes sense because lower income households are likely to be more sensitive to price and perceived value for money offered by fast-food stores, and may also rely more on small, frequent shopping trips close to home rather than large weekly shopping trips to a large supermarket further away, and on the convenience of food outlets near home^31^.

Unlike for fast-food proximity and availability of formal PA facilities, there was little evidence of an association between neighbourhood greenspace – operationalised as domestic gardens and public greenspace with 300 m of home – and CVD-related hospital admissions for any income or deprivation subgroup. Instead, the null association observed for the whole sample was largely preserved across household income and area deprivation levels, with no evidence of effect modification. The observation that area deprivation drives a positive departure from additivity for formal PA facilities but not for greenspace may reflect poorer quality greenspace^32^ or perceived safety concerns in deprived areas^16^, or other factors making greenspaces less suitable for PA. It is worth noting that the number of observations in the most deprived, most green combination was small, leading to a lack of precision around that hazard ratio, and correspondingly around the RERI. Thus, we should be cautious about drawing conclusions about whether area deprivation modifies the estimated effect of greenspace on CVD risk.

### Strengths and limitations

This study demonstrates the value of examining effect modification by variables such as income and area deprivation, when a study is sufficiently powered to do so, to better understand potential equity impacts of interventions, and to avoid overlooking potential health benefits for some groups even when population-wide effects may be small or negligible. It is highly plausible that some demographic groups will exhibit greater sensitivity to their neighbourhood environment than others, and that some groups may not be affected at all. Estimating average effects across the whole population smooths out these differences and potentially leads to erroneous conclusions about the importance of neighbourhood environments for certain subpopulations. Our findings also demonstrate the value of examining effect modification by multiple socioeconomic indicators in parallel and in combination – as this can yield important insights that may otherwise be missed when we focus only on a single measure of either household or area-level socioeconomic conditions.

The study has several limitations. First, the hospital admissions data only captures inpatient care, so any early detection of CVD that occurs in primary care settings after baseline and is then effectively treated without admission to hospital will not be counted. Such cases are probably more likely to occur in higher income or less deprived subgroups^33^, and this may have contributed to lower risk of the outcome in those groups, potentially distorting the magnitude of effect modification on the additive scale. Linkage of GP records to the UK Biobank cohort should make it possible to examine this potential source of bias in future. Related to this, if some types of health care have shifted to outpatient settings over the course of the follow-up period, some dilution of the true association overall and between subgroups may have occurred.

Second, it is unclear what period of follow-up is necessary to capture the effect of interest, given that people will have been exposed to their baseline neighbourhood conditions for varying lengths of time depending on how long they have lived at that address, whether relevant changes had occurred in their neighbourhood during that time, and the nature of previous neighbourhood exposures. We adjusted our analyses for years living at baseline address to attempt to deal with this and are reassured by the lengthy average time people have lived at the same address (median=15 years), but there may be remaining imprecision, and potential bias of estimates in either direction, that we cannot overcome using observational data of this kind. Longer follow up may prove to be more revealing, and that will become possible in future years, but ideally future work would also account for changes in the built environment over that period. UK Biobank would be made richer by the addition of measurement of neighbourhood exposures at one or more post-baseline time points. Our sensitivity analyses using a shorter follow-up period to account for the timing of the exposure ascertainment showed that most point estimates were robust to this specification, but there was a loss of precision presumably driven by the substantial reduction in the number of hospital admissions occurring during the shortened follow-up period (Supplementary Table 2).

Third, there is likely to be some systematic misclassification, random error, and geographical inconsistency in quality in the measure of the neighbourhood environment we have used, particularly the fats-food and physical activity environment measures, due to our reliance on ‘off-the-shelf’ measures based on local authority data sources collected for non-research purposes. Relatedly, the food environment measure captures only proximity and not the density of outlets in an area, and nor does it take into account the wider neighbourhood food environment, in terms of access to both healthy and unhealthy food, which are often highly correlated^34^. The greenspace measure may also not adequately capture the full extent of relevant greenness of one’s neighbourhood, as it does not include smaller parcels of greenspace such a street trees or reflect ‘quality’ of greenspace.

Finally, we cannot rule out self-selection into more health promoting neighbourhoods by people more disposed to healthy behaviours. We can, however, by the longitudinal nature of the study and exclusion of people with prevalent disease at baseline, rule out active self-selection prior to baseline into neighbourhoods on the basis of prevalent disease (e.g. following a cardiac event earlier in life, deciding to relocate to a neighbourhood more supportive of a healthy lifestyle). This means that we likely minimise masking of the true effect via this avenue, but may still have some residual positive confounding that could bias the association away from the null, despite our comprehensive adjustment for observed potential confounders. However, UK Biobank is a residentially very stable sample, and most of our strongest findings were within more deprived subgroups, where financial resources enabling relocation for health purposes are presumably the least.

## Conclusions

Greater availability of PA facilities close to home is associated with lower risk of CVD admissions in more deprived areas, but only among those with higher household incomes. Improving deprived neighbourhoods by increasing the number of formal PA facilities, while also ensuring access to these is free or affordable, and by limiting the proximity of fast-food outlets to residential areas, may improve health outcomes in more deprived areas. Examining effect modification by multiple socioeconomic indicators can potentially uncover important associations that would otherwise be masked and contribute to our understanding of the multiple ways in which people’s socioeconomic conditions influence their relationship with the built environment and its effects on their health. Such insights can help better inform the design and targeting of neighbourhood built environment interventions in a way that avoids generating or perpetuating health inequalities.

## Data Availability

All data used in this study are available to approved researchers on application to UK Biobank https://www.ukbiobank.ac.uk/

