## Supplementary material for "Neighbourhood built environments, socioeconomic position, and hospital admissions for cardiovascular disease: a prospective study using UK Biobank"

### 1. Primary analysis stratified by sex

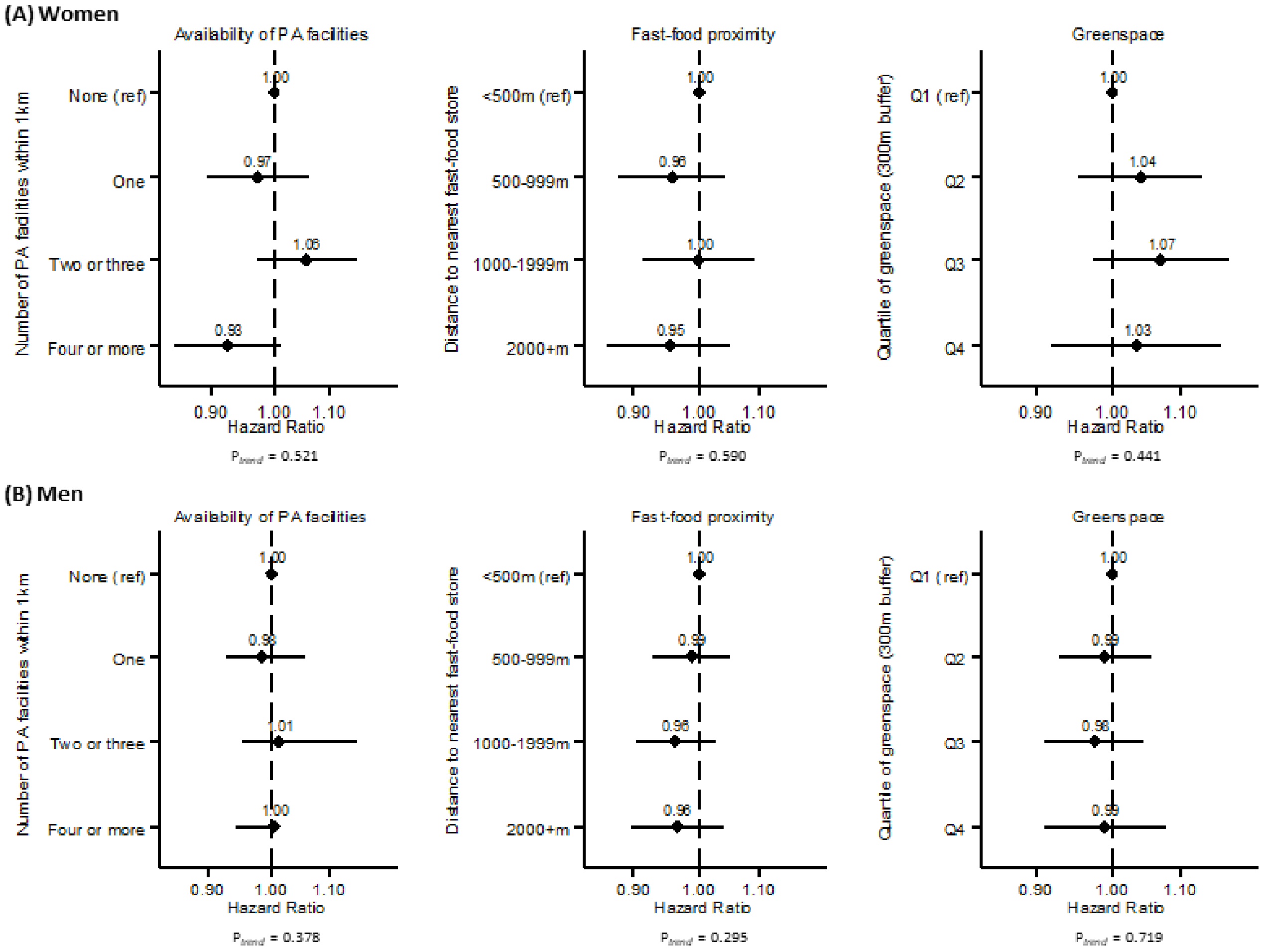


**Supplementary Figure 1: Hazard ratios for associations between neighbourhood characteristics and CVD-related hospital admissions, stratified by sex**

**Supplementary Table 1a. Modification of the association between neighbourhood availability of PA facilities and hospital admissions due to CVD, by household income and area deprivation, stratified by sex**

| | *Annual household income* | | | | *Area deprivation* | | | |
|---|---|---|---|---|---|---|---|---|
| | **WOMEN** | | **MEN** | | **WOMEN** | | **MEN** | |
| | **< £31,000** | **At least £31,000** | **< £31,000** | **At least £31,000** | **More deprived** | **Less deprived** | **More deprived** | **Less deprived** |
| **Number of PA facilities** | HR (95% CI); P | HR (95% CI); P | HR (95% CI); P | HR (95% CI); P | HR (95% CI); P | HR (95% CI); P | HR (95% CI); P | HR (95% CI); P |
| None | 1.00 (ref) | 0.92 (0.82, 1.03) P=0.165 | 1.00 (ref) | 1.01 (0.93, 1.09) P=0.814 | 1.00 (ref) | 0.87 (0.76, 1.00) P=0.046 | 1.00 (ref) | 0.89 (0.80, 0.98) P=0.015 |
| One | 0.96 (0.87, 1.06) P=0.435 | 0.91 (0.80, 1.05) P=0.196 | 1.03 (0.95, 1.11) P=0.539 | 0.94 (0.86, 1.04) P=0.222 | 0.93 (0.79, 1.10) P=0.415 | 0.86 (0.75, 0.99) P=0.038 | 0.93 (0.82, 1.05) P=0.235 | 0.89 (0.80, 0.98) P=0.020 |
| 2-3 | 1.05 (0.95, 1.15) P=0.353 | 0.98 (0.87, 1.12) P=0.809 | 1.03 (0.95, 1.11) P=0.449 | 1.00 (0.91, 1.09) P=0.968 | 1.09 (0.94, 1.26) P=0.261 | 0.91 (0.79, 1.04) P=0.160 | 0.93 (0.83, 1.04) P=0.179 | 0.93 (0.84, 1.02) P=0.138 |
| 4 or more | 0.93 (0.84, 1.03) P=0.167 | 0.83 (0.73, 0.95) P=0.006 | 1.05 (0.97, 1.14) P=0.248 | 0.95 (0.87, 1.04) P=0.286 | 0.89 (0.77, 1.04) P=0.145 | 0.83 (0.72, 0.95) P=0.009 | 0.93 (0.83, 1.04) P=0.181 | 0.92 (0.83, 1.02) P=0.095 |
| Stratum-specific HRs (4+ facilities vs 0) | 0.94 (0.84, 1.05) P=0.245 | 0.88 (0.76, 1.04) P=0.129 | 1.04 (0.96, 1.14) P=0.327 | 0.94 (0.86, 1.04) P=0.227 | 0.92 (0.78, 1.09) P=0.334 | 0.94 (0.85, 1.05) P=0.308 | 0.89 (0.79, 1.01) P=0.064 | 1.04 (0.96, 1.12) P=0.315 |
| Relative excess risk due to interaction (RERI) | -0.018 (-0.168, 0.132) P=0.813 | | -0.105 (-0.218, 0.009) P=0.070 | | 0.061 (-0.096, 0.218) P=0.446 | | 0.102 (-0.012, 0.217) P=0.081 | |

**Supplementary Table 1b. Modification of the association between fast-food proximity and hospital admissions due to CVD, by household income and area deprivation, stratified by sex**

| | *Annual household income* | | | | *Area deprivation* | | | |
|---|---|---|---|---|---|---|---|---|
| | **WOMEN** | | **MEN** | | **WOMEN** | | **MEN** | |
| | **< £31,000** | **At least £31,000** | **< £31,000** | **At least £31,000** | **More deprived** | **Less deprived** | **More deprived** | **Less deprived** |
| **Fast-food proximity** | HR (95% CI); P | HR (95% CI); P | HR (95% CI); P | HR (95% CI); P | HR (95% CI); P | HR (95% CI); P | HR (95% CI); P | HR (95% CI); P |
| Closer than 500m | 1.00 (ref) | 0.96 (0.83, 1.11) P=0.609 | 1.00 (ref) | 0.90 (0.81, 0.99) P=0.039 | 1.00 (ref) | 0.93 (0.81, 1.05) P=0.246 | 1.00 (ref) | 1.01 (0.92, 1.11) P=0.851 |
| 500-999m | 0.97 (0.88, 1.07) P=0.539 | 0.89 (0.78, 1.01) P=0.079 | 0.97 (0.90, 1.05) P=0.521 | 0.91 (0.83, 1.00) P=0.044 | 0.99 (0.88, 1.13) P=0.934 | 0.85 (0.76, 0.96) P=0.009 | 1.02 (0.93, 1.12) P=0.661 | 0.96 (0.88, 1.05) P=0.391 |
| 1000-1999m | 1.03 (0.93, 1.14) P=0.584 | 0.90 (0.79, 1.02) P=0.111 | 0.93 (0.86, 1.01) P=0.087 | 0.91 (0.83, 1.00) P=0.044 | 0.99 (0.86, 1.14) P=0.889 | 0.91 (0.81, 1.03) P=0.120 | 0.96 (0.87, 1.07) P=0.479 | 0.95 (0.87, 1.03) P=0.225 |
| At least 2000m | 0.95 (0.85, 1.07) P=0.395 | 0.91 (0.79, 1.04) P=0.171 | 0.92 (0.85, 1.01) P=0.086 | 0.92 (0.84, 1.01) P=0.091 | 1.01 (0.86, 1.20) P=0.868 | 0.85 (0.74, 0.97) P=0.014 | 1.03 (0.91, 1.17) P=0.616 | 0.94 (0.85, 1.03) P=0.168 |
| Stratum-specific HRs (≥2000m vs <500m) | 0.93 (0.82, 1.05) P=0.226 | 0.99 (0.82, 1.18) P=0.880 | 0.92 (0.84, 1.02) P=0.107 | 1.02 (0.91, 1.14) P=0.722 | 0.98 (0.82, 1.18) P=0.857 | 0.92 (0.81, 1.04) P=0.201 | 1.06 (0.92, 1.22) P=0.394 | 0.92 (0.84, 1.00) P=0.057 |
| Relative excess risk due to interaction (RERI) | -0.007 (-0.183, 0.170) P=0.942 | | 0.098 (-0.017, 0.213) P=0.095 | | -0.092 (-0.290, 0.106) P=0.361 | | -0.107 (-0.259, 0.046) P=0.169 | |

**Supplementary Table 1c. Modification of the association between neighbourhood greenspace and hospital admissions due to CVD, by household income and area deprivation, stratified by sex**

| | *Annual household income* | | | | *Area deprivation* | | | |
|---|---|---|---|---|---|---|---|---|
| | **WOMEN** | | **MEN** | | **WOMEN** | | **MEN** | |
| | **< £31,000** | **At least £31,000** | **< £31,000** | **At least £31,000** | **More deprived** | **Less deprived** | **More deprived** | **Less deprived** |
| **Greenspace** | HR (95% CI); P | HR (95% CI); P | HR (95% CI); P | HR (95% CI); P | HR (95% CI); P | HR (95% CI); P | HR (95% CI); P | HR (95% CI); P |
| Q1 (least greenspace) | 1.00 (ref) | 0.91 (0.79, 1.04) P=0.154 | 1.00 (ref) | 0.91 (0.83, 1.00) P=0.046 | 1.00 (ref) | 0.86 (0.75, 0.97) P=0.016 | 1.00 (ref) | 1.05 (0.95, 1.15) P=0.339 |
| Q2 | 1.03 (0.93, 1.14) P=0.608 | 0.97 (0.85, 1.11) P=0.702 | 0.98 (0.90, 1.06) P=0.598 | 0.92 (0.83, 1.01) P=0.067 | 1.01 (0.89, 1.14) P=0.874 | 0.90 (0.81, 1.00) P=0.057 | 1.03 (0.94, 1.13) P=0.576 | 0.97 (0.89, 1.05) P=0.389 |
| Q3 | 1.06 (0.95, 1.18) P=0.315 | 0.99 (0.87, 1.14) P=0.941 | 0.95 (0.88, 1.04) P=0.278 | 0.92 (0.84, 1.01) P=0.083 | 1.04 (0.89, 1.21) P=0.621 | 0.92 (0.82, 1.02) P=0.116 | 1.05 (0.93, 1.17) P=0.446 | 0.94 (0.87, 1.02) P=0.154 |
| Q4 (most greenspace) | 1.04 (0.91, 1.18) P=0.587 | 0.93 (0.80, 1.08) P=0.330 | 0.95 (0.86, 1.05) P=0.276 | 0.95 (0.86, 1.05) P=0.331 | 0.97 (0.79, 1.19) P=0.759 | 0.89 (0.78, 1.01) P=0.070 | 0.98 (0.84, 1.15) P=0.847 | 0.97 (0.88, 1.06) P=0.497 |
| Stratum-specific HRs (Q4 vs Q1) | 1.03 (0.89, 1.19) P=0.691 | 1.03 (0.84, 1.27) P=0.758 | 0.97 (0.86, 1.08) P=0.565 | 1.01 (0.89, 1.15) P=0.859 | 0.86 (0.67, 1.11) P=0.244 | 1.06 (0.91, 1.22) P=0.456 | 1.02 (0.84, 0.23) P=0.878 | 0.91 (0.82. 1.01) P=0.077 |
| Relative excess risk due to interaction (RERI) | -0.015 (-0.186, 0.155) P=0.861 | | 0.096 (-0.017, 0.208) P=0.095 | | 0.063 (-0.155, 0.280) P=0.573 | | -0.062 (-0.233, 0.109) P=0.475 | |

### 2. Sensitivity analyses: Restricting follow-up time to January 2012 onwards

#### Supplementary Table 3. Hospital admissions by household income and area deprivation (follow-up time restricted to January 2012 onwards)

| | CVD | |
|---|---|---|
| | N | Admissions (%) |
| Total | 328030 | 7698 (2.3) |
| Household income (annual pre-tax) | | |
| <£31,000 | 152833 | 4497 (2.9) |
| £31,000 or more | 175197 | 3201 (1.8) |
| Area deprivation | | |
| More deprived | 94488 | 2342 (2.5) |
| Less deprived | 233542 | 5356 (2.3) |

**Supplementary Table 4. Modification of the associations between neighbourhood environment variables and hospital admissions due to CVD, by household income and area deprivation (follow-up time restricted to January 2012 onwards)**

| | *Annual household income* | | *Area deprivation* | |
|---|---|---|---|---|
| | **Less than £31,000** | **At least £31,000** | **More deprived** | **Less deprived** |
| **Number of PA facilities** | HR (95% CI); P | HR (95% CI); P | HR (95% CI): P | HR (95% CI); P |
| None | 1.00 (ref) | 0.99 (0.92, 1.10) P=0.934 | 1.00 (ref) | 0.89 (0.80, 0.98) P=0.024 |
| One | 1.00 (0.91, 1.08) P=0.852 | 0.94 (0.88, 1.08) P=0.577 | 0.89 (0.77, 1.01) P=0.077 | 0.90 (0.81, 1.01) P=0.061 |
| 2-3 | 1.04 (0.96, 1.13) P=0.290 | 0.99 (0.92, 1.12) P=0.727 | 1.01 (0.90, 1.14) P=0.852 | 0.92 (0.82, 1.02) P=0.119 |
| 4 or more | 1.00 (0.95, 1.13) P=0.393 | 0.92 (0.84, 1.02) P=0.133 | 0.94 (0.84, 1.06) P=0.326 | 0.90 (0.80, 1.00) P=0.058 |
| Stratum-specific HRs (4+ facilities vs 0) | 1.03 (0.95, 1.13) P=0.466 | 0.92 (0.83, 1.02) P=0.130 | 0.94 (0.83, 1.07) P=0.354 | 1.01 (0.93, 1.10) P=0.772 |
| Relative excess risk due to interaction (RERI) | -0.113 (-0.236, 0.010) P=0.072 | | 0.070 (-0.056, 0.197) P=0.274 | |
| **Fast-food proximity** | HR (95% CI); P | HR (95% CI); P | HR (95% CI): P | HR (95% CI); P |
| Closer than 500m | 1.00 (ref) | 0.90 (0.80, 1.00) P=0.059 | 1.00 (ref) | 0.98 (0.88, 1.08) P=0.667 |
| 500-999m | 0.94 (0.86, 1.02) P=0.141 | 0.91 (0.82, 1.00) P=0.051 | 1.01 (0.91, 1.12) P=0.864 | 0.90 (0.82, 1.00) P=0.039 |
| 1000-1999m | 0.95 (0.87, 1.04) P=0.247 | 0.93 (0.85, 1.03) P=0.173 | 0.98 (0.88, 1.10) P=0.766 | 0.94 (0.86, 1.03) P=0.207 |
| At least 2000m | 0.94 (0.85, 1.03) P=0.182 | 0.92 (0.83, 1.02) P=0.132 | 1.00 (0.87, 1.15) P=0.977 | 0.92 (0.83, 1.02) P=0.118 |
| Stratum-specific HRs (≥2000m vs <500m) | 0.94 (0.85, 1.04) P=0.243 | 1.02 (0.90, 1.16) P=0.725 | 0.99 (0.85, 1.15) P=0.874 | 0.95 (0.86, 1.04) P=0.263 |
| Relative excess risk due to interaction (RERI) | 0.087 (-0.040, 0.213) P=0.179 | | -0.058 (-0.217, 0.101) P=0.476 | |
| **Greenspace** | HR (95% CI); P | HR (95% CI); P | HR (95% CI): P | HR (95% CI); P |
| Q1 (least greenspace) | 1.00 (ref) | 0.91 (0.83, 1.01) P=0.090 | 1.00 (ref) | 0.92 (0.84, 1.02) P=0.118 |
| Q2 | 1.00 (0.92, 1.08) P=0.932 | 0.94 (0.85, 1.04) P=0.209 | 0.97 (0.88, 1.08) P=0.599 | 0.94 (0.86, 1.02) P=0.134 |
| Q3 | 0.97 (0.89, 1.07) P=0.576 | 0.99 (0.90, 1.10) P=0.892 | 1.03 (0.91, 1.17) P=0.610 | 0.93 (0.85, 1.01) P=0.088 |
| Q4 (most greenspace) | 0.98 (0.88, 1.09) P=0.717 | 0.95 (0.84, 1.06) P=0.338 | 0.94 (0.80, 1.12) P=0.499 | 0.92 (0.83, 1.02) P=0.120 |
| Stratum-specific HRs (Q4 vs Q1) | 0.99 (0.88, 1.12) P=0.908 | 1.02 (0.89, 1.18) P=0.738 | 0.88 (0.72, 1.08) P=0.228 | 1.01 (0.90, 1.12) P=0.925 |
| Relative excess risk due to interaction (RERI) | 0.050 (-0.076, 0.176) P=0.435 | | 0.056 (-0.119, 0.231) P=0.530 | |

**Supplementary Table 5. Association between neighbourhood characteristics and CVD-related hospital admissions, stratified by household income and area deprivation in combination (follow-up time restricted to January 2012 onwards)**

| | *Combined household income and area deprivation* | | | |
|---|---|---|---|---|
| | **Less than £31,000 & more deprived** | **At least £31,000 & more deprived** | **Less than £31,000 & less deprived** | **At least £31,000 & less deprived** |
| | HR (95% CI); P | HR (95% CI); P | HR (95% CI); P | HR (95% CI); P |
| **Number of PA facilities** | | | | |
| None | 1.00 (ref) | 1.00 (ref) | 1.00 (ref) | 1.00 (ref) |
| One | 0.95 (0.81, 1.11) P=0.524 | 0.75 (0.57, 0.97) P=0.030 | 1.01 (0.91, 1.13) P=0.789 | 1.01 (0.90, 1.13) P=0.847 |
| 2-3 | 1.08 (0.93, 1.25) P=0.338 | 0.85 (0.68, 1.07) P=0.174 | 1.03 (0.92, 1.14) P=0.627 | 1.03 (0.92, 1.15) P=0.611 |
| 4 or more | 1.04 (0.89, 1.22) P=0.593 | 0.73 (0.57, 0.92) P=0.008 | 1.03 (0.92, 1.16) P=0.572 | 0.97 (0.86, 1.10) P=0.659 |
| $P_{trend}$ | 0.335 | 0.030 | 0.541 | 0.809 |
| | HR (95% CI); P | HR (95% CI); P | HR (95% CI); P | HR (95% CI); P |
| **Fast-food proximity** | | | | |
| Closer than 500m | 1.00 (ref) | 1.00 (ref) | 1.00 (ref) | 1.00 (ref) |
| 500-999m | 1.01 (0.89, 1.14) P=0.894 | 1.03 (0.85, 1.24) P=0.795 | 0.89 (0.79, 1.00) P=0.053 | 0.99 (0.86, 1.14) P=0.864 |
| 1000-1999m | 0.95 (0.83, 1.09) P=0.483 | 1.08 (0.87, 1.34) P=0.493 | 0.95 (0.84, 1.07) P=0.401 | 1.00 (0.87, 1.15) P=0.977 |
| At least 2000m | 0.93 (0.78, 1.11) P=0.425 | 1.15 (0.87, 1.52) P=0.334 | 0.94 (0.82, 1.06) P=0.311 | 0.99 (0.85, 1.15) P=0.866 |
| $P_{trend}$ | 0.342 | 0.310 | 0.704 | 0.920 |
| | HR (95% CI); P | HR (95% CI); P | HR (95% CI); P | HR (95% CI); P |
| **Greenspace** | | | | |
| Q1 (least greenspace) | 1.00 (ref) | 1.00 (ref) | 1.00 (ref) | 1.00 (ref) |
| Q2 | 0.94 (0.83, 1.06) P=0.313 | 1.08 (0.89, 1.31) P=0.425 | 1.04 (0.92, 1.18) P=0.535 | 1.00 (0.86, 1.16) P=0.993 |
| Q3 | 1.03 (0.88, 1.20) P=0.695 | 0.98 (0.76, 1.28) P=0.901 | 0.97 (0.85, 1.11) P=0.649 | 1.10 (0.95, 1.28) P=0.211 |
| Q4 (most greenspace) | 0.92 (0.72, 1.17) P=0.494 | 0.81 (0.54, 1.20) P=0.289 | 1.02 (0.87, 1.19) P=0.804 | 1.05 (0.89, 1.25) P=0.543 |
| $P_{trend}$ | 0.799 | 0.571 | 0.797 | 0.321 |

### 3. Sensitivity analyses: Models additionally adjusted for baseline BMI, hypertension and medications for hypertension and high cholesterol

#### Supplementary Table 6. Modification of the association between neighbourhood environment variables and hospital admissions due to CVD, by household income and area deprivation (adjusted for additional risk factors)

| | *Annual household income* | | *Area deprivation* | |
|---|---|---|---|---|
| | **Less than £31,000** | **At least £31,000** | **More deprived** | **Less deprived** |
| **Number of PA facilities** | HR (95% CI); P | HR (95% CI); P | HR (95% CI): P | HR (95% CI); P |
| None | 1.00 (ref) | 0.99 (0.93, 1.06) P=0.730 | 1.00 (ref) | 0.91 (0.85, 0.99) P=0.028 |
| One | 1.00 (0.95, 1.08) P=0.793 | 0.94 (0.87, 1.01) P=0.100 | 0.93 (0.85, 1.03) P=0.191 | 0.91 (0.84, 0.99) P=0.036 |
| 2-3 | 1.04 (0.98, 1.11) P=0.140 | 0.99 (0.93, 1.07) P=0.998 | 0.99 (0.91, 1.09) P=0.912 | 0.96 (0.88, 1.04) P=0.305 |
| 4 or more | 1.00 (0.97, 1.10) P=0.371 | 0.92 (0.86, 1.00) P=0.046 | 0.95 (0.87, 1.04) P=0.240 | 0.93 (0.85, 1.01) P=0.087 |
| Stratum-specific HRs (4+ facilities vs 0) | 1.02 (0.96, 1.10) P=0.479 | 0.94 (0.87, 1.02) P=0.140 | 0.93 (0.84, 1.03) P=0.149 | 1.02 (0.96, 1.09) P=0.531 |
| Relative excess risk due to interaction (RERI) | -0.090 (-0.182, 0.002) P=0.055 | | 0.066 (-0.030, 0.163) P=0.177 | |
| **Fast-food proximity** | HR (95% CI); P | HR (95% CI); P | HR (95% CI): P | HR (95% CI); P |
| Closer than 500m | 1.00 (ref) | 0.91 (0.84, 0.99) P=0.031 | 1.00 (ref) | 1.00 (0.92, 1.08) P=0.957 |
| 500-999m | 0.97 (0.91, 1.03) P=0.343 | 0.91 (0.84, 0.98) P=0.009 | 1.01 (0.93, 1.09) P=0.849 | 0.95 (0.88, 1.02) P=0.133 |
| 1000-1999m | 0.97 (0.91, 1.03) P=0.346 | 0.91 (0.84, 0.98) P=0.013 | 0.97 (0.89, 1.06) P=0.500 | 0.96 (0.89, 1.03) P=0.260 |
| At least 2000m | 0.93 (0.87, 1.00) P=0.050 | 0.92 (0.85, 1.00) P=0.049 | 1.02 (0.92, 1.13) P=0.657 | 0.93 (0.86, 1.00) P=0.064 |
| Stratum-specific HRs (≥2000m vs <500m) | 0.93 (0.86, 1.00) P=0.051 | 1.01 (0.92, 1.12) P=0.767 | 1.04 (0.93, 1.16) P=0.498 | 0.92 (0.86, 0.99) P=0.030 |
| Relative excess risk due to interaction (RERI) | 0.081 (-0.015, 0.177) P=0.098 | | -0.092 (-0.214, 0.030) P=0.141 | |
| **Greenspace** | HR (95% CI); P | HR (95% CI); P | HR (95% CI): P | HR (95% CI); P |
| Q1 (least greenspace) | 1.00 (ref) | 0.91 (0.84, 0.98) P=0.015 | 1.00 (ref) | 0.99 (0.92, 1.07) P=0.879 |
| Q2 | 0.99 (0.93, 1.05) P=0.640 | 0.93 (0.86, 1.00) P=0.058 | 1.01 (0.94, 1.09) P=0.778 | 0.96 (0.90, 1.02) P=0.197 |
| Q3 | 0.99 (0.92, 1.06) P=0.714 | 0.94 (0.87, 1.01) P=0.089 | 1.03 (0.94, 1.13) P=0.489 | 0.96 (0.89, 1.02) P=0.171 |
| Q4 (most greenspace) | 0.97 (0.90, 1.05) P=0.515 | 0.95 (0.88, 1.04) P=0.266 | 0.97 (0.85, 1.10) P=0.611 | 0.97 (0.90, 1.04) P=0.401 |
| Stratum-specific HRs (Q4 vs Q1) | 0.99 (0.91, 1.09) P=0.883 | 1.02 (0.91, 1.13) P=0.774 | 0.96 (0.82, 1.12) P=0.596 | 0.97 (0.89, 1.05) P=0.409 |
| Relative excess risk due to interaction (RERI) | 0.071 (-0.023, 0.165) P=0.141 | | 0.006 (-0.129, 0.141) P=0.934 | |

**Supplementary Table 7. Association between neighbourhood characteristics and CVD-related hospital admissions, stratified by household income and area deprivation in combination (adjusted for additional risk factors)**

| | *Combined household income and area deprivation* | | | |
|---|---|---|---|---|
| | **Less than £31,000 & more deprived** | **At least £31,000 & more deprived** | **Less than £31,000 & less deprived** | **At least £31,000 & less deprived** |
| | HR (95% CI); P | HR (95% CI); P | HR (95% CI): P | HR (95% CI); P |
| **Number of PA facilities** | | | | |
| None | 1.00 (ref) | 1.00 (ref) | 1.00 (ref) | 1.00 (ref) |
| One | 0.97 (0.86, 1.09) P=0.629 | 0.82 (0.66, 1.00) P=0.050 | 1.03 (0.95, 1.11) P=0.527 | 0.97 (0.89, 1.05) P=0.468 |
| 2-3 | 1.01 (0.90, 1.13) P=0.880 | 0.91 (0.76, 1.09) P=0.293 | 1.07 (0.99, 1.15) P=0.109 | 1.02 (0.94, 1.11) P=0.665 |
| 4 or more | 0.99 (0.88, 1.11) P=0.846 | 0.79 (0.65, 0.95) P=0.011 | 1.04 (0.96, 1.14) P=0.330 | 0.98 (0.89, 1.07) P=0.645 |
| $P_{trend}$ | 0.997 | 0.035 | 0.192 | 0.886 |
| | HR (95% CI); P | HR (95% CI); P | HR (95% CI): P | HR (95% CI); P |
| **Fast-food proximity** | | | | |
| Closer than 500m | 1.00 (ref) | 1.00 (ref) | 1.00 (ref) | 1.00 (ref) |
| 500-999m | 1.02 (0.93, 1.12) P=0.614 | 1.00 (0.86, 1.16) P=0.991 | 0.93 (0.85, 1.01) P=0.101 | 0.98 (0.88, 1.09) P=0.732 |
| 1000-1999m | 0.96 (0.87, 1.07) P=0.467 | 1.05 (0.88, 1.24) P=0.587 | 0.96 (0.88, 1.05) P=0.366 | 0.97 (0.87, 1.08) P=0.557 |
| At least 2000m | 1.00 (0.88, 1.14) P=0.998 | 1.16 (0.94, 1.44) P=0.171 | 0.89 (0.81, 0.98) P=0.018 | 0.98 (0.88, 1.10) P=0.767 |
| $P_{trend}$ | 0.678 | 0.209 | 0.055 | 0.786 |
| | HR (95% CI); P | HR (95% CI); P | HR (95% CI): P | HR (95% CI); P |
| **Greenspace** | | | | |
| Q1 (least greenspace) | 1.00 (ref) | 1.00 (ref) | 1.00 (ref) | 1.00 (ref) |
| Q2 | 1.00 (0.91, 1.10) P=0.956 | 1.10 (0.95, 1.28) P=0.217 | 0.99 (0.90, 1.08) P=0.753 | 0.95 (0.85, 1.07) P=0.409 |
| Q3 | 1.04 (0.93, 1.17) P=0.479 | 1.05 (0.86, 1.29) P=0.632 | 0.98 (0.88, 1.08) P=0.618 | 0.97 (0.87, 1.09) P=0.622 |
| Q4 (most greenspace) | 0.92 (0.77, 1.11) P=0.381 | 1.10 (0.83, 1.47) P=0.499 | 0.99 (0.89, 1.11) P=0.913 | 0.98 (0.87, 1.12) P=0.807 |
| $P_{trend}$ | 0.937 | 0.404 | 0.893 | 0.950 |

### 4. Examination of proportional hazards assumption

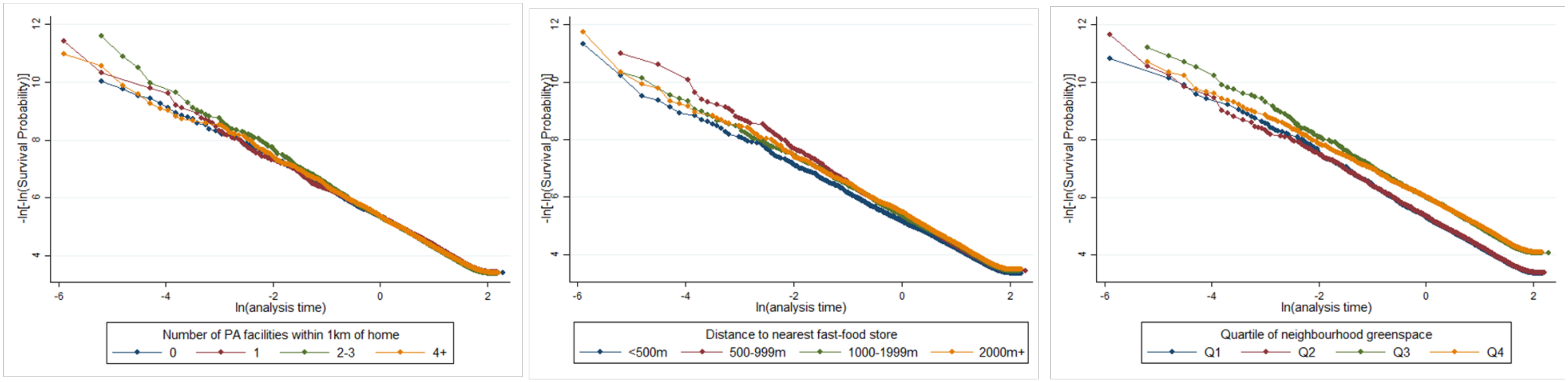


**Supplementary Figure 2. Log-log plots (adjusted for all covariates) for graphical examination of proportional hazards assumption**
